# Prevalence of metabolic disorders in obese triple negative breast cancer patients

**DOI:** 10.1101/2020.05.07.20094037

**Authors:** Luz Angela Torres-de la Roche, Alina Jara Schulte, Rajesh Devassy, Harald Krentel, Kay Willborn, Jennifer Eidswick, Rudy Leon De Wilde

## Abstract

**Introduction:** Obesity is a risk factor for hormone receptor positive breast cancer in postmenopausal women. However, the association between triple negative breast cancer risk and metabolic abnormalities is not yet clear. Moreover, little is known regarding the prevalence of these abnormalities in this group of women. Here we present the prevalence of metabolic abnormalities in a single cohort of obese postmenopausal triple negative breast cancer patients.

**Methods:** Monocentric, retrospective, single cohort analysis of triple negative breast cancer patients treated between January 2008 and December 2017 at Pius Hospital Oldenburg. For quantitative or numerical variables, central tendency and dispersion measures were used. Values are presented as mean number of patients or percentage.

**Results:** Among 2745 breast cancer cases, 43 obese postmenopausal patients with a mean age of 64 years (range 51 to 90) had triple negative breast cancer. Most were diagnosed with invasive ductal (n = 39; 90.7 %), high-grade carcinoma (n = 35; 81.4 %), with a tumor size between 2 and 5 cm (n = 19; 44.2 %). Half the cohort lacked lymph node involvement; 5 patients showed distant metastasis (11.6 %). The majority had no family history of breast cancer (n = 32; 74.4 %), were non-smokers (n = 37; 86 %), and had a history of pregnancy (n = 35; 81.4 %). Frequent metabolic abnormalities included hypertension (n = 31; 72.1 %) and dyslipidemia (n = 36; 83.7 %) whereas type 2 diabetes or glucose intolerance were less prevalent (n = 13; 30.2 %).

**Conclusion:** Hypertension and dyslipidemia were more prevalent in the study cohort than type 2 diabetes. Moderately obese patients were most frequently affected. These findings partially align with international studies which observed an association between triple negative breast cancer and elevated levels of blood glucose and triglycerides, but not between tumor disease and hypertension.

## Introduction

Breast cancer (BC) is the second most common cancer type worldwide,[1]. In 2018, two million new cases of BC occurred in women worldwide, and more than 600,000 female patients died due to BC,[2]. In western countries such as Germany, BC is the most commonly diagnosed tumor disease and the leading cause of cancer-related death in women. Cancer registry data published by the Robert Koch-Institute (RKI) show that in Germany in 2014, 69,220 women were newly diagnosed with BC (incidence rate 114.6/100,000) and 17,670 deaths in women were caused by BC (mortality rate 23.0/100,000),[3]. Obesity, defined as a BMI ≥ 30 kg/m^2^, increases the overall risk of BC 1.5-fold in postmenopausal women,[4]. As a serious medical condition with increased mortality, it contributes greatly to the development of multiple chronic diseases, such as arteriosclerosis, diabetes, and cancer,[5]. According to RKI in 2014, 53 % of all women in Germany were considered overweight, and 24 % even suffered from severe obesity,[6]. The steadily rising prevalence of obesity and associated secondary diseases results in extreme costs for the social system, making these topics public health issues of high relevance [7].

Triple negative receptor breast cancer (TNBC) is a molecular subtype of BC defined by a lack of estrogen receptor (ER), progesterone receptor (PR), and HER2 expression,[8] Approximately 10-20 % of BC patients are diagnosed with TNBC, with relatively young women frequently affected,[9]. It exhibits aggressive clinical behavior, with a high rate of recurrence and distant metastasis resulting in poor clinical outcome,[10]. In a 2014 meta-analysis, Bhandari et al. observed a positive association between metabolic syndrome (MS) and risk of BC, which increased when cases were limited to postmenopausal patients,[11]. Moreover, a prospective study by Park et al. demonstrated that women who fell within the normal weight range (> 18 kg/m^2^< 25 kg/m^2^) and who exhibited one or more metabolic abnormalities were at an increased risk of developing BC, whereas women who fell within the overweight (< 25 kg/m^2^) to obese (≥30 kg/m^2^) range had an increased risk of postmenopausal BC seemingly regardless of concomitant metabolic abnormalities,[12]. Nevertheless, the association between metabolic abnormalities and risk of TNBC in obese postmenopausal women remains unclear,[13-15]. In particular, there is a lack of evidence regarding the prevalence of metabolic abnormalities among TNBC patients with different levels of obesity.

We analyzed the oncological outcomes of 2,745 BC patients treated between January 2008 and December 2017, at Pius Hospital Oldenburg, Germany. Here we present the prevalence of the metabolic abnormalities in the subgroup of postmenopausal patients diagnosed with TNBC according to different categories of obesity (low-grade obesity: BMI of 30 to 34.9 kg/m^2^, moderate obesity: BMI of 35 to 39.9 kg/m^2^, high-grade obesity: BMI of ≥ 40 kg/m^2^).

## Methods

Monocentric, transversal, descriptive study of a single cohort of patients treated for BC between January 2008 and December 2017, at Pius Hospital Oldenburg, were selected according to the International Classification of Diseases (ICD-10) from the hospital’s Tumor Documentation database. Medical records were accessed via Orbis, the database program of Pius Hospital Oldenburg. Anonymized case data were collected in SPSS (software version 25.0).

Suitable cases were selected according to the following inclusion criteria: female patient, confirmed diagnosis of TNBC, postmenopausal status, and a BMI of ≥ 30 kg/m^2^. Cases of carcinoma in situ of the breast, relapse, ER, PR and/or HER2 positive status, premenopausal and perimenopausal status, and/or normal weight or pre-obesity (BMI of ≤ 29.9 kg/m^2^), as well as those for whom variables of interest were missing were excluded.

For statistical analysis, patient characteristics, such as age and risk factors for BC (early menarche, late menopause, nulliparity, no breastfeeding), as well as tumor characteristics, including histopathological features and stage according to TNM classification, were collected from clinical charts. Variables were analyzed by means of descriptive statistics. For quantitative or numerical variables, central tendency measures (mean, median, mode), and dispersion measures (standard deviation, or ranges) were used. Arithmetic mean (%) was used for qualitative variables.

In order to analyze the levels of obesity, the study population was subdivided into three groups of different BMI categories. Group 1 included postmenopausal TNBC patients with low-grade obesity (BMI between 30 and 34.9 kg/m^2^), group 2 consisted of postmenopausal TNBC patients with moderate obesity (BMI between 35 and 39.9 kg/m^2^), and group 3 represented postmenopausal TNBC patients with high-grade obesity (BMI of ≥ 40 kg/m^2^).

Prior to study initiation, approval from the IRB of Pius Hospital and the Ethics committee of Carl von Ossietzky University was obtained (2018/139; 07.01.2019; 05.02.2019).

## Results

Between January 2008 and December 2017, 2745 cases of BC were diagnosed and documented at Pius Hospital Oldenburg (Figure 1). After filtering for TNBC cases, the list of 279 TNBC patients was subdivided into three stages of menopause from which only the postmenopausal patients (n = 191) were retained for analysis and subsequently filtered for patients with a BMI of ≥ 30 kg/m^2^. The selection process resulted in a study population of 43 obese patients. Group 1 (low-grade obesity) consisted of 29 patients (67.44 %). Group 2 (moderate obesity) and Group 3 (high-grade obesity) each included 7 patients (16.28 % respectively).

The study population (Table 1) consisted of 43 obese, postmenopausal patients with a mean age of 64 years (SD ± 9.5). The greatest proportion (n = 29; 67.44 %) had a BMI between 30 and 34.9 kg/m^2^, followed by 7 patients (16.28 %) with a BMI between 35 and 39.9 kg/m^2^, and another 7 (16.28 %) with a BMI ≥ 40 kg/m^2^. A large majority (n=39; 90.7 %) had been diagnosed with invasive ductal TNBC, followed by 2 patients (4.7 %) with neuroendocrine TNBC. Inflammatory TNBC and spindle cell type were each represented by 1 patient (2.3 %).

The evaluation of the tumor size according to TNM revealed that the majority of these patients (n = 19; 44.2 %) had been diagnosed with TNBC sized between 2 cm and 5 cm, closely followed by 16 patients (37.2 %) with a tumor size smaller than 2 cm. Five patients (11.6 %) accounted for TNBC larger than 5 cm and 3 patients (7 %) had tumors that extended to the chest wall or skin. Lymph nodes of 24 patients (55.8 %) were not affected by the tumor, whereas 11 patients (25.6 %) were diagnosed with involvement of movable axillary lymph nodes level I-II. Three patients (7 %) had involvement of fixed axillary lymph nodes level I-II, and 4 patients (9.3 %) showed infiltration of supra- or infraclavicular lymph nodes level III.

**Table 1:** Clinical characteristics at initial diagnosis of obese patients with TNBC

| **Variables** |  |  |
| --- | --- | --- |
| **Age (years)** | Mean: 64,47 (SD: ± 9.5) | Range: 51 – 90 |
|  | **Total (n = 43)** | **%** |
| **BMI (kg/m<sup>2</sup>)** |  |  |
| 30-34.9 | 29 | 67.44 |
| 35-39.9 | 7 | 16.28 |
| $\geq 40$ | 7 | 16.28 |
| **Histopathology** |  |  |
| Invasive ductal | 39 | 90.7 |
| Inflammatory | 1 | 2.3 |
| Neuroendocrine | 2 | 4.7 |
| Spindle cell | 1 | 2.3 |
| **Tumor size** |  |  |
| T1 | 16 | 37.2 |
| T2 | 19 | 44.2 |
| T3 | 5 | 11.6 |
| T4 | 3 | 7 |
| **Lymph node involvement** |  |  |
| N0 | 24 | 55.8 |
| N1 | 11 | 25.6 |
| N2 | 3 | 7 |
| N3 | 4 | 9.3 |
| Unknown | 1 | 2.3 |
| **Distant metastasis** |  |  |
| M0 | 38 | 88.4 |
| M1 | 5 | 11.6 |
| **Grading** |  |  |
| G2 | 8 | 18.6 |
| G3 | 35 | 81.4 |
| **Family history of BC** |  |  |
| Positive | 11 | 25.6 |
| Negative | 32 | 74.4 |
| **Smoking status** |  |  |
| Active smoker | 6 | 14 |
| Non-smoker | 32 | 74.4 |
| Former smoker | 5 | 11.6 |
| **Gravidity** |  |  |
| Positive | 35 | 81.4 |
| Negative | 8 | 18.6 |
| **Hypertension** |  |  |
| Positive | 31 | 72.1 |
| Negative | 12 | 27.9 |
| **Diabetes mellitus II** |  |  |
| Positive | 8 | 18.6 |
| Negative | 30 | 69.8 |
| Glucose intolerance | 5 | 11.6 |
| **Dyslipidemia** |  |  |
| Positive | 36 | 83.7 |
| Negative | 7 | 16.3 |

Most of the patients (38; 88.4 %) were not affected by distant metastasis, however in 5 patients (11.6 %) the tumor had already metastasized at time of diagnosis. Grading results showed that the majority (n = 35; 81.4 %) had high-grade TNBC in contrast to 8 patients (18.6 %) with moderate grade.

Data regarding family history of BC revealed that 11 patients (25.6 %) had at least one first degree relative with BC. However, the majority (n = 32; 74.4 %) had a negative family history of BC. The analysis of further risk factors for BC showed that the most (n = 32; 74.4 %) had never smoked, whilst 6 (14 %) were active smokers at time of diagnosis, and 5 (11.6 %) had given up smoking prior to diagnosis. Furthermore, the majority of the study population (n = 35; 81.4 %) had been pregnant at least once in their lifetime. Data on the number of pregnancies as well as breastfeeding history was incomplete and, therefore, excluded from the analysis.

With regard to metabolic abnormalities, most patients could be characterized as having dyslipidemia (n = 36; 83.7 %) and/or hypertension (n = 31; 72.1 %). However, only 13 patients (30.2 %) were diagnosed with type 2 diabetes or showed signs of glucose intolerance.

According to the different BMI categories (Table 2), Group 1 had the highest mean age (66 years vs. 59 years in Group 2 vs. 64 years in Group 3). Group 3 presented the largest age range with the youngest patient being 54 and the oldest 90 years old at time of diagnosis. Whereas all patients in Group 2 and 3 were diagnosed with invasive ductal TNBC, patients with rare BC subtypes were seen only in Group 1: two patients (6.9 %) with neuroendocrine TNBC, one case (3.4 %) of inflammatory TNBC, and one patient with spindle cell type TNBC (3.4 %).

In all three groups, most of the patients were diagnosed with TNBC sized 5 cm or less (79.3 % in Group 1, 100 % in Group 2, 71.5 % in Group 3). Group 3 presented the highest percentage of patients with tumors that had already extended to the chest wall at time of diagnosis (n = 3; 28.6 %). Group 1 included six patients (20.6 %) with TNBC larger than 5 cm, among which one patient (3.4 %) was diagnosed with tumor expansion to the chest wall.

**Table 2:** Clinical characteristics of obese patients with TNBC according to BMI

| Variables |  | Group 1<br>n = 29 |  | Group 2<br>n = 7 |  | Group 3<br>n = 7 |
| --- | --- | --- | --- | --- | --- | --- |
| Age (years) |  |  |  |  |  |  |
| Mean |  | 65.69 |  | 59.43 |  | 64.43 |
| SD |  | ± 9.2 |  | ± 6 |  | ± 12.7 |
| Range |  | 53 – 87 |  | 51 – 67 |  | 54 – 90 |
| Histopathology |  |  |  |  |  |  |
| Invasive ductal | 25 | 82.8 % | 7 | 100 % | 7 | 100 % |
| Inflammatory | 1 | 3.4 % | 0 | 0 % | 0 | 0 % |
| Neuroendocrine | 2 | 6.9 % | 0 | 0 % | 0 | 0 % |
| Spindle cell | 1 | 3.4 % | 0 | 0 % | 0 | 0 % |
| Tumor size |  |  |  |  |  |  |
| T1 | 11 | 37.9 % | 3 | 42.9 % | 2 | 28.6 % |
| T2 | 12 | 41.4 % | 4 | 57.1 % | 3 | 42.9 % |
| T3 | 5 | 17.2 % | 0 | 0 % | 0 | 0 % |
| T4 | 1 | 3.4 % | 0 | 0 % | 3 | 28.6 % |
| Lymph node involvement |  |  |  |  |  |  |
| N0 | 15 | 51.7 % | 6 | 85.7 % | 3 | 42.9 % |
| N1 | 8 | 27.6 % | 1 | 14.3 % | 2 | 28.6 % |
| N2 | 2 | 6.9 % | 0 | 0 % | 1 | 14.3 % |
| N3 | 4 | 13.8 % | 0 | 0 % | 0 | 0 % |
| Unknown | 0 | 0 % | 0 | 0 % | 1 | 14.3 % |
| Distant metastasis |  |  |  |  |  |  |
| M0 | 24 | 82.8 % | 7 | 100 % | 7 | 100 % |
| M1 | 5 | 17.2 % | 0 | 0 % | 0 | 0 % |
| Grading |  |  |  |  |  |  |
| G2 | 6 | 20.7 % | 2 | 28.6 % | 0 | 0 % |
| G3 | 23 | 79.3 % | 5 | 71.4 % | 7 | 100 % |
| Family history |  |  |  |  |  |  |
| Positive | 8 | 27.6 % | 1 | 14.3 % | 2 | 28.6 % |
| Negative | 21 | 72.4 % | 6 | 85.7 % | 5 | 71.4 % |
| Smoking status |  |  |  |  |  |  |
| Active smoker | 4 | 13.8 % | 1 | 14.3 % | 1 | 14.3 % |
| Non-smoker | 20 | 69 % | 6 | 85.7 % | 6 | 85.7 % |
| Former smoker | 5 | 17.2 % | 0 | 0 % | 0 | 0 % |
| Gravidity |  |  |  |  |  |  |
| Positive | 25 | 86.2 % | 4 | 57.1 % | 6 | 85.7 % |
| Negative | 4 | 13.8 % | 3 | 42.9 % | 1 | 14.3 % |
| Hypertension |  |  |  |  |  |  |
| Positive | 21 | 72.4 % | 6 | 85.7 % | 4 | 57.1 % |
| Negative | 8 | 27.6 % | 1 | 14.3 % | 3 | 42.9 % |
| Diabetes mellitus II |  |  |  |  |  |  |
| Positive | 2 | 6.9 % | 3 | 42.9 % | 3 | 42.9 % |
| Negative | 22 | 75.9 % | 4 | 57.1 % | 4 | 57.1 % |
| Glucose intolerance | 5 | 17.2 % | 0 | 0 % | 0 | 0 % |
| Dyslipidemia |  |  |  |  |  |  |
| Positive | 25 | 86.2 % | 6 | 85.7 % | 5 | 71.4 % |
| Negative | 4 | 13.8 % | 1 | 14.3 % | 2 | 28.6 % |

Most cases across the groups showed no lymph node involvement at the time of diagnosis (51.7 % in Group 1, 85.7 % in Group 2, 42.9 % in Group 3). Group 1 and 3 had a similar percentage of patients with infiltration of movable axillary lymph nodes level I-II (27.6 % in Group 1, 28.6 % in Group 3). However, involvement of supra- and/or infraclavicular lymph nodes level III appeared only in Group 1 (n = 4; 13.8 %). In addition, solely Group 1 included patients with distant metastasis at time of diagnosis (n= 5; 17.2 %).

The majority of patients in all three groups were diagnosed with high-grade TNBC (79.3 % in Group 1; 71.4 % in Group 2, 100 % in Group 3). Group 3 proved to be exclusively high-grade, whereas Group 1 and Group 2 presented similar percentages of patients with moderate TNBC (20.7 % in Group 1; 28.6 % in Group 2).

Compared to Group 2, the percentage of patients with a family history positive for BC in Groups 1 and 3 was twice as high (27.6 % in Group 1 and 28.6 % in Group 3 compared to 14.3 % in Group 2).

Groups 2 and 3 each included six non-smoking patients (85.7 %) and one active smoker (14.3 %). In Group 1, 69 % were non-smokers compared to 13.8 % active smokers and 17.2 % former smokers. Whilst almost half of the patients in Group 2 reported zero past pregnancies, only 14 % of Groups 1 and 3 were nulligravida.

The analysis of metabolic abnormalities revealed that Group 1 and Group 2 had higher percentages of patients with hypertension and dyslipidemia than those with high-grade obesity (Group 3). Whilst approximately 70 % of all patients in Group 1 and more than 85 % in Group 2 suffered from elevated blood pressure levels, only half of the patients in Group 3 had been diagnosed with hypertension. In addition, more than 85 % both in Groups 1 and 2 showed signs of dyslipidemia, whereas 70 % of Group 3 had lipid metabolism disorders. A diagnosis of type 2 diabetes was seen in 40 % of both Groups 2 and 3. In comparison, Group 1 had a three-quarter majority of patients without signs of glucose intolerance.

Taking all results into account, the group with low-grade obesity presented histopathological tumor features associated with a poorer prognosis compared to patients in Group 2 and 3 with higher grades of obesity. In addition, patients in Group 1 exhibited BC risk factors more often than Groups 2 and 3. Group 2 with moderate obesity was most frequently affected by metabolic abnormalities, whereas the group with high-grade obesity presented the lowest percentage.

## Discussion

The association between TNBC and metabolic abnormalities has not yet been elucidated in German studies and international studies on this topic lack differentiation between various grades of obesity and require further research. The selection process in this study resulted in 279 TNBC cases among 2745 registered BC patients between January 2008 and December 2017, at Pius Hospital. Though located at the very bottom of the range, the percentage of 10.16 % TNBC cases is in line with various sources which state that this molecular subtype makes up between 10 and 20 % of all BC cases,[8,9,10,16].

With a mean age of 64 years at time of diagnosis (SD ± 9.5), the study population agrees with epidemiological surveys which determined 64 years to be the average age of BC onset,[7]. However, when comparing to epidemiological data on TNBC only, our study population is ten years older than average,[17-18]. In our study, two thirds of the patient population exhibited low-grade obesity (BMI 30-34.9 kg/m^2^). Group 2, representing moderate obesity, is the only subgroup in which patients were comparatively younger when diagnosed with BC (59 years; SD ± 6), although still older than in comparable studies.

Data on histopathological features revealed that most of the patients in the study population were diagnosed with high-grade, invasive ductal TNBC sized larger than 2 cm. Fifty percent showed no signs of lymph node involvement, and only five patients with low-grade obesity had distant metastasis at time of diagnosis. These findings are in accordance with results from a Dutch study conducted by Kreike and colleagues,[19]. Among their 97 TNBC cases, 87 % were high-grade and 83 % invasive ductal carcinomas. Sixty-five percent had TNBC sized larger than 2 cm, and 56 % were not affected by lymph node involvement.

Statistical analysis for established risk factors showed that most of our cohort did not have a familial predisposition for BC, active smoking status, and/or nulliparity. In terms of these factors, the moderately obese group had the most unfavorable risk profile for BC and the low-grade obesity group presented more clinical features associated with a poorer prognosis compared to patients in the more obese group. Regarding risk factors, a case-control study by Ma et al. which included 554 TNBC (among 2,658 BC cases) stated that established risk factors for BC might not apply in unrestricted manners to TNBC,[20]. In their case-control studies of 2,658 BC patients aged 20-64 years, they found that age at menarche, age at first full-term pregnancy, and nulliparity were not associated with risk of TNBC, while TNBC risk decreased with an increasing duration of breastfeeding.

The analysis of metabolic abnormalities revealed that the majority of our study population suffered from hypertension and/or dyslipidemia, but type 2 diabetes and/or glucose intolerance were less prevalent. Regarding all three metabolic abnormalities, those with moderate obesity were most frequently affected, whereas those with high-grade obesity yielded the lowest frequency.

Our findings differ from results of international studies on the association of metabolic abnormalities and TNBC. In a prospective cohort study including 22,833 female BC patients, Yang et al. investigated the association between blood pressure and risk of BC overall according to molecular subtype. Their results supported previous findings that hypertension has no impact on risk of BC,[21]. Maiti and colleagues came to a similar conclusion in their retrospective study examining the association between MS and TNBC which included 86 TNBC patients (among 176 BC cases). However, whereas hypertension and BMI > 25 kg/m^2^ were not associated with TNBC, blood glucose level > 100 mg/dl, HDL cholesterol level < 50 mg/dl, and triglyceride level > 150 mg/dl were associated. Moreover, Maiti et al. observed a significantly increased prevalence of MS in TNBC patients and supported the hypothesis that TNBC cases present a higher histological tumor grade,[18].

Due to the lack of evidence about the prevalence of TNBC among different levels of obesity, we could not compare our findings. The strength of this retrospective study is the large cohort of BC patients analyzed. However, since information on waist circumference was not documented on all medical charts, central obesity could not be determined with absolute certainty. Therefore, the study population could not be examined for MS and comparisons to other studies were limited to statements about separate components of MS.

## Conclusion

In summary, our findings are partially in accordance with international studies which have reported an association between TNBC and elevated levels of blood glucose and triglycerides, but not between TNBC and hypertension. According to our results, obese TNBC patients present at initial diagnosis with different metabolic abnormalities, with hypertension and dyslipidemia being the most prevalent, especially in women of moderate obesity. The group with high-grade obesity showed a lower frequency of hypertension and dyslipidemia but a higher prevalence of type 2 diabetes. Further studies should clarify which metabolic abnormalities are prevalent in BC patients and which lead to increased risk of TNBC. A deeper understanding of how each different metabolic disorder influences the development of TNBC could supply important information for the establishment of new prevention and treatment concepts. Meanwhile, we recommend physicians provide women personalized management of obesity and metabolic disorders to reduce their risk of developing this highly aggressive form of cancer.

## Data Availability

The anonymized data base could be transferred to regulatory bodies on request

## Disclaimers

The views expressed in the submitted article are our own and not an official position of the institutions where the authors work. We the authors declare that this manuscript is original, has not been published before and is not currently being considered for publication elsewhere.

We confirm that the manuscript has been read and approved by all named authors and that there are no other persons who satisfied the criteria for authorship but are not listed. We further confirm that the order of authors listed in the manuscript has been approved by all of us.

We understand that the Corresponding Author, Prof. Dr. Dr. med. Rudy Leon De Wilde, is the sole contact for the Editorial process (including Editorial Manager and direct communications with the office). He is responsible for communicating with the other authors about progress, submissions of revisions and final approval of proofs, and had listed in the Acknowledgments everyone who contributed significantly to the study.

We confirm that we have provided a current, correct email address which is accessible by the Corresponding Author and which has been configured to accept email from. Phone: (49) (+ 441) 229 1524

## Supplementary material

None.

